# The E-Patient Quality Improvement and Standardization (EQIS) Platform for Improving Maternal and Pediatric Clinical Care in Bangladesh: Study Protocol for a Prospective Open Cohort Study

**DOI:** 10.64898/2026.01.07.26343582

**Authors:** Salahuddin Ahmed, Ahad Mahmud Khan, Sifat Binte Ibrahim, Farhana Ahmad, Md. Minhajul Hossain, Anindita Tabassum, Sumaiya Afroze Khan, Sabbir Ahmed, Nabidul Haque Chowdhury, Md. Shafiqul Islam, Chowdhury Ali Kawser, Mohammod Shahidullah, Ferdousi Begum, Firoza Begum, Farhana Dewan, Steven Johnson, Othman Ouenes, Mohammad Elias, Abdullah H. Baqui, John Peabody

**Affiliations:** Projahnmo Research Foundation, Dhaka, Bangladesh; Bangladesh College of Physicians and Surgeons, Dhaka, Bangladesh; Obstetrical and Gynecological Society of Bangladesh, Dhaka, Bangladesh; Peabody Health Philanthropy, San Francisco, CA, USA; Elias & Sultana Foundation, Windsor Mill, MD, USA; Department of International Health, Johns Hopkins Bloomberg School of Public Health, Baltimore, Maryland, USA; School of Medicine, University of California, San Francisco, CA, USA; School of Pubic Health, University of California, Los Angeles, CA, USA

## Abstract

**Introduction:** Inappropriate healthcare practices are major contributors to poor outcomes of maternal and pediatric patients. Where resources are scarce, provider-level targeted interventions, such as implementation of standardized, evidence-based protocols and quality improvement initiatives are particularly needed to address these gaps in care. Simulation-based tools have been shown to have a promising effect on improving clinical care management. This study aims to evaluate the effectiveness of a simulation-based platform called E-Patient Quality Improvement and Standardization (EQIS) in improving maternal and pediatric health care in Bangladesh.

**Methods and analysis:** The study employs a prospective, open-cohort design and will be conducted across five tertiary-level hospitals in Dhaka, Bangladesh. All physicians engaged in providing maternal or pediatric care will be recruited. The EQIS platform will facilitate the development of online simulated cases, aiming to enhance physician practice and patient outcomes through involvement and immediate personalized feedback. The development of cases will consist of several stages of review by expert clinicians, ensuring that the cases align with national and local practice guidelines. Each participant physician will work on three cases per round, completing one of six rounds every four months for two years. Data will be collected and analyzed after each round. Descriptive and inferential analysis will be done to assess trends in clinical performance and pinpoint areas for improvement. In addition, patient-level pre-and post-impact data will be collected. These data will evaluate the impact of the EQIS platform on physician performance, as measured by changes in patient satisfaction, treatment costs, referrals, and facility utilization.

**Ethics and dissemination:** The study protocol was approved by the National Research Ethics Committee of the Bangladesh Medical Research Council (registration number: 59524062024). Dissemination of the study findings will be through meetings with stakeholders, conference presentations and publications in peer-reviewed journals.

**Strengths and limitations of this study:**

- This study will use simulated cases for the first time in Bangladesh to improve physicians’ standardization in diagnosing and treating maternal and pediatric patients.
- We postulate that the EQIS platform will help physicians to improve and standardize their critical thinking and decision-making skills by presenting a variety of clinical scenarios. With this platform, physicians will improve their skills in diagnosing and treating various medical conditions, in accordance with national and international guidelines, without endangering patient safety.
- This platform will provide an opportunity for physicians to receive prompt, serial feedback and self-reflection. This will help them learn from mistakes and possibly improve their clinical capability.
- The study will be carried out in five tertiary-level hospitals in Dhaka city, Bangladesh, including one public hospital, one autonomous hospital, and three private hospitals. This approach might not represent the broader population or reflect healthcare practices in rural areas and lower-tier facilities across the country.

## INTRODUCTION

Maternal and child mortality are major global health issues. In 2023, there were an estimated 260,000 maternal deaths and 4.8 million deaths of children under five. Most of these deaths occurred in low- and middle-income countries (LMICs).^1–3^ Bangladesh is a clear example of this crisis, showing a maternal mortality ratio of 156 per 100,000 live births^4^ and an under-five mortality rate of 31 per 1,000 live births^5^. Scientific evidence highlights several preventable causes of maternal and child mortality. Hemorrhage, hypertensive disorders, infections, and complications from pre-existing medical conditions are among the leading causes of maternal deaths,^6–8^ while neonatal conditions such as preterm birth complications, intrapartum-related events, neonatal sepsis, respiratory infections, diarrheal diseases, and malnutrition dominate as the primary contributors to under-five mortality.^9^

A critical, yet often underappreciated, contributor to these outcomes is the inappropriate quality of clinical care.^10^ ^11^ Inadequate clinical practices, particularly in maternal and pediatric services, can result in catastrophic consequences for patients.^12^ ^13^ These practices are driven by multiple interrelated factors, including insufficient training, poor adherence to evidence-based guidelines, lack of continuous professional development, and systemic health system barriers such as understaffing and high patient loads.^14–18^ Furthermore, significant variability in clinical decision-making, where patients with similar conditions receive different treatments depending on the provider, further exacerbates disparities in care.^19^ ^20^

Improving maternal and pediatric health outcomes in resource-limited settings requires the implementation of standardized, evidence-based clinical practices supported by quality improvement initiatives.^21^ Applying clinical guidelines during critical periods has shown significant potential to improve care quality and reduce mortality.^22^ Quality improvement initiatives play a crucial role in this process by addressing gaps in clinical practice and ensuring high standards of patient care.^23^ Practicing evidence-based care, strengthening health infrastructure and enhancing the skills of healthcare providers ultimately contributes to improved patient outcomes and reduced disparities.^21^ ^22^ ^24^

Among these strategies, simulation-based tools have proven highly effective in enhancing clinical performance by providing healthcare providers with risk-free, practical learning environments ^25–27^. These platforms enable the safe exploration of complex clinical scenarios. They help to build providers’ confidence, clinical reasoning, and decision-making skills.^25^ Evidence from studies assessing simulation-based tools in healthcare show they effectively improve both clinical performance and patient outcomes. Several studies have reported positive impacts of simulation-based training on healthcare learners’ knowledge, clinical competencies and patient level outcomes.^27–32^, while other studies reported no significant difference in skill acquisition and performance in the clinical practice environment after using this tool in LMIC’s.^33^

The E-Patient Quality Improvement and Standardization (EQIS) platform and its widely used commercial predecessor CPV^®^s are recognized, as an effective resource aimed at standardizing care and improving patient outcomes^31^ ^34^ ^35^. EQIS is a systematic and structured way to assess, evaluate, and improve clinical performance based on established standards, guidelines, and benchmarks. EQIS simulations present healthcare providers with a diverse array of clinical scenarios, including rare and complex cases. This helps them develop critical thinking and decision-making skills.^36^ The platform provides immediate feedback, enabling providers to spot and fix errors, sharpen their clinical judgment, and improve their performance. Ultimately, this method improves clinical skills and self-confidence, leading to better outcomes in real-world settings.^25^ ^27^ ^37^

A study by Peabody et al. across six low-income countries showed significant improvements in adherence to clinical guidelines, reduced practical variation, and enhanced patient safety after using the EQIS platform.^38^ In other LMIC settings, this platform has been used to identify differences in practice patterns across healthcare sites and among providers with varying levels of training and experience. Providers who received simulation-based feedback demonstrate consistent improvements in performance scores over time, which correlated with greater adherence to clinical guidelines, shorter hospital stays, and substantial cost savings.^39^

The quality of clinical care practices in hospitals across Bangladesh is often reported to be below average.^40–43^ While the EQIS platform has demonstrated effectiveness in various healthcare settings, its impact in severely resource-constrained environments like Bangladesh remains underexplored and underutilized. This study protocol details an initiative to evaluate how the EQIS platform can improve and standardize care practices for mothers and children in Bangladesh. Through rigorous assessment, the study aims to contribute to the existing evidence that supports simulation-based quality improvement interventions in LMICs and to provide practical insights for health systems working to meet the Sustainable Development Goals (SDGs) related to maternal and child health.

### Objectives

1. To measure the effectiveness of the EQIS platform in improving the quality of clinical performance among physicians in Obstetrics/Gynecological and Pediatric departments.
2. To measure the effectiveness of the EQIS platform in standardizing clinical practices among physicians in those departments.

## METHODS AND ANALYSIS

### Study design and settings

This will be a prospective, open-cohort study. The study will be conducted in the obstetrics and gynecological and pediatric departments of five purposively selected hospitals in Dhaka, Bangladesh (Figure 1): Shaheed Suhrawardy Medical College and Hospital (ShSMCH), Ashulia Women and Children Hospital (AWCH), Institute of Child and Mother Health (ICMH), Dr. MR Khan Shishu Hospital, and Obstetrical and Gynecological Society of Bangladesh and Hospital (OGSBH). These facilities represent a mix of one public, one autonomous, and three private institutions. This diversity in healthcare delivery models will provide valuable insights into the usability of the EQIS platform and its effectiveness in standardizing and improving maternal and pediatric clinical care across different settings.

**Figure 1:**
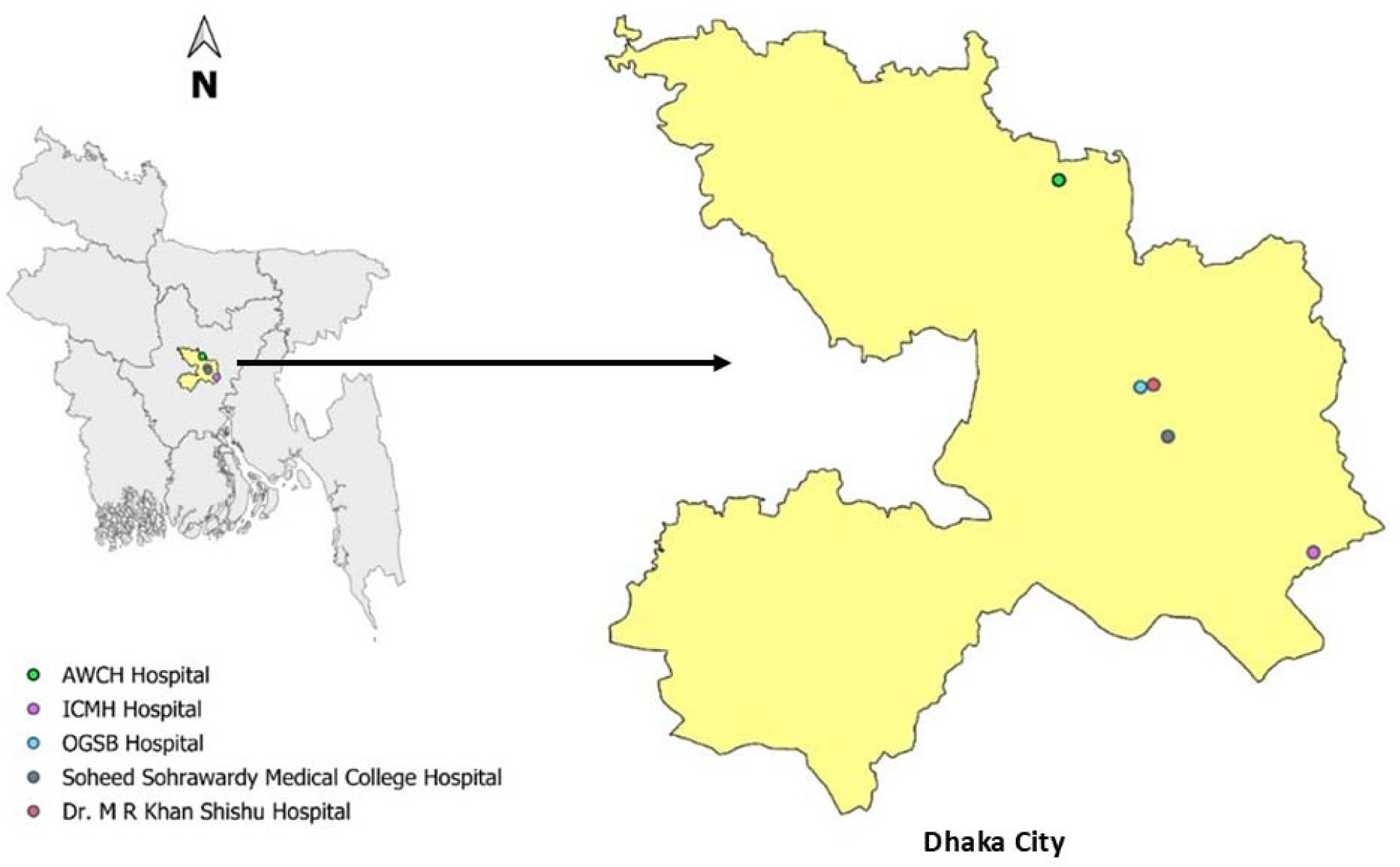
Study hospitals in Dhaka city.

The study will be conducted over a period of three years, consisting of three phases: (i) a six-month preparatory phase that includes case development and listing of physicians; (ii) a two-year implementation phase during which the EQIS platform will be applied across six evaluation points—history taking, physical examination, laboratory and imaging work-up, diagnostic accuracy, treatment, and follow up— to assess the physicians’ clinical performance and (iii) a six-month phase for data analysis, report writing, and dissemination. The study started in August 2024 and is planned to be completed by July 2027.

### Study population

The study population will be physicians in the Obstetrics and Gynecology department and Pediatric departments of the selected hospitals. Physicians working in those departments and providing care to the patients will be included. We will also enroll physicians who will join the hospital during the study period. Physicians will be excluded if they are not directly involved in patient care during the study period – this will include those on extended leave, those who have transferred to other departments or institutions, or those who have shifted their field of clinical practice. In addition, medical students and intern doctors will be excluded to ensure that all participants have a requisite level of experience and clinical decision-making responsibility.

### Study procedure

The study procedure has been presented in **Figure 2**.

**Figure 2:**
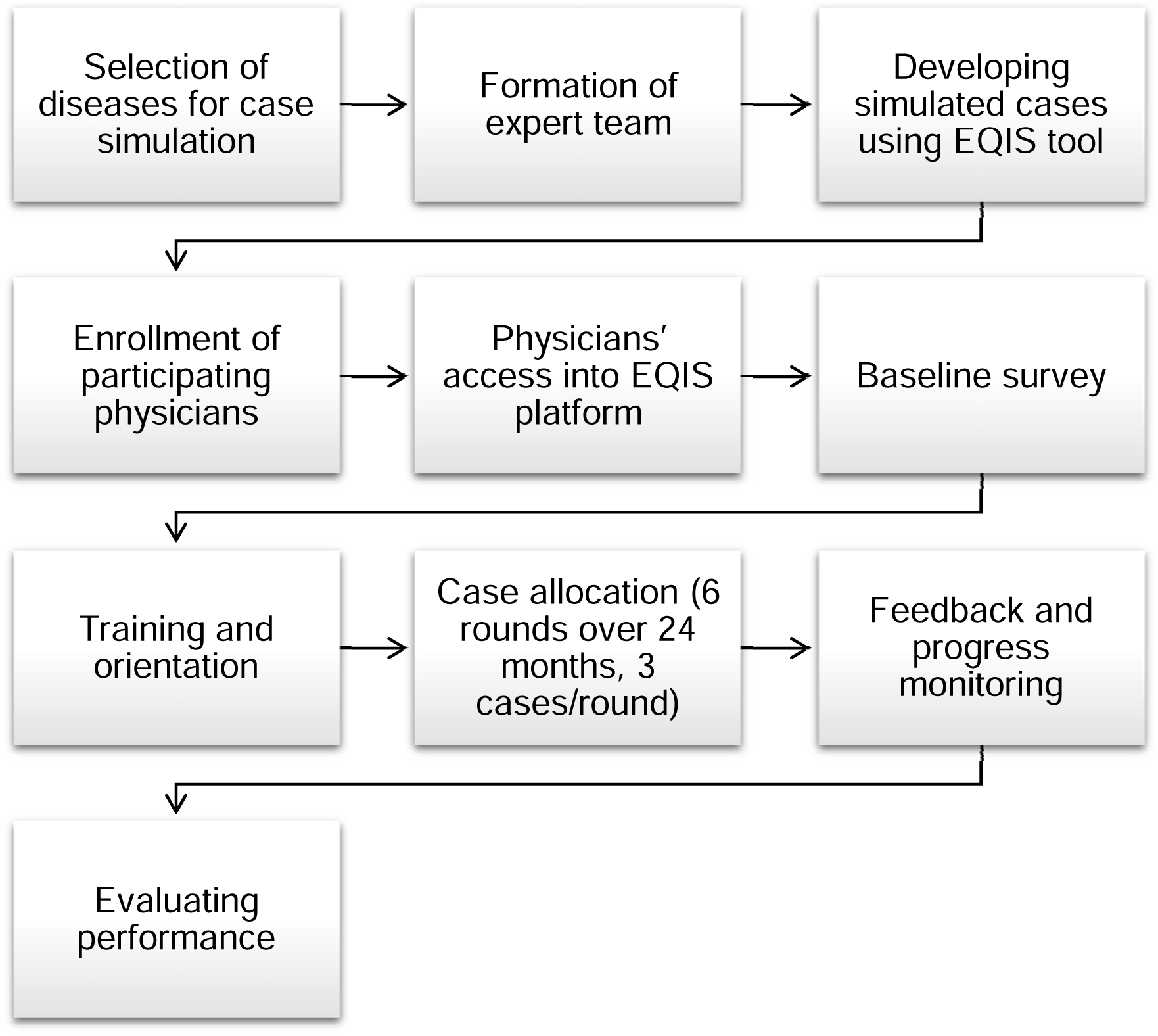
Study procedure.

#### Selection of diseases for case simulation

The research team has already been formed. They reviewed Bangladesh specific literature and discussed with obstetricians, gynecologists, pediatricians from several hospitals, and representatives of professional bodies to identify the three leading causes of maternal and pediatric morbidity and mortality. Based on this, the team finalized three maternal and three pediatric topics, each comprising six separate cases—totaling 18 cases for maternal and 18 for pediatric care (Table 1).

**Table 1:**
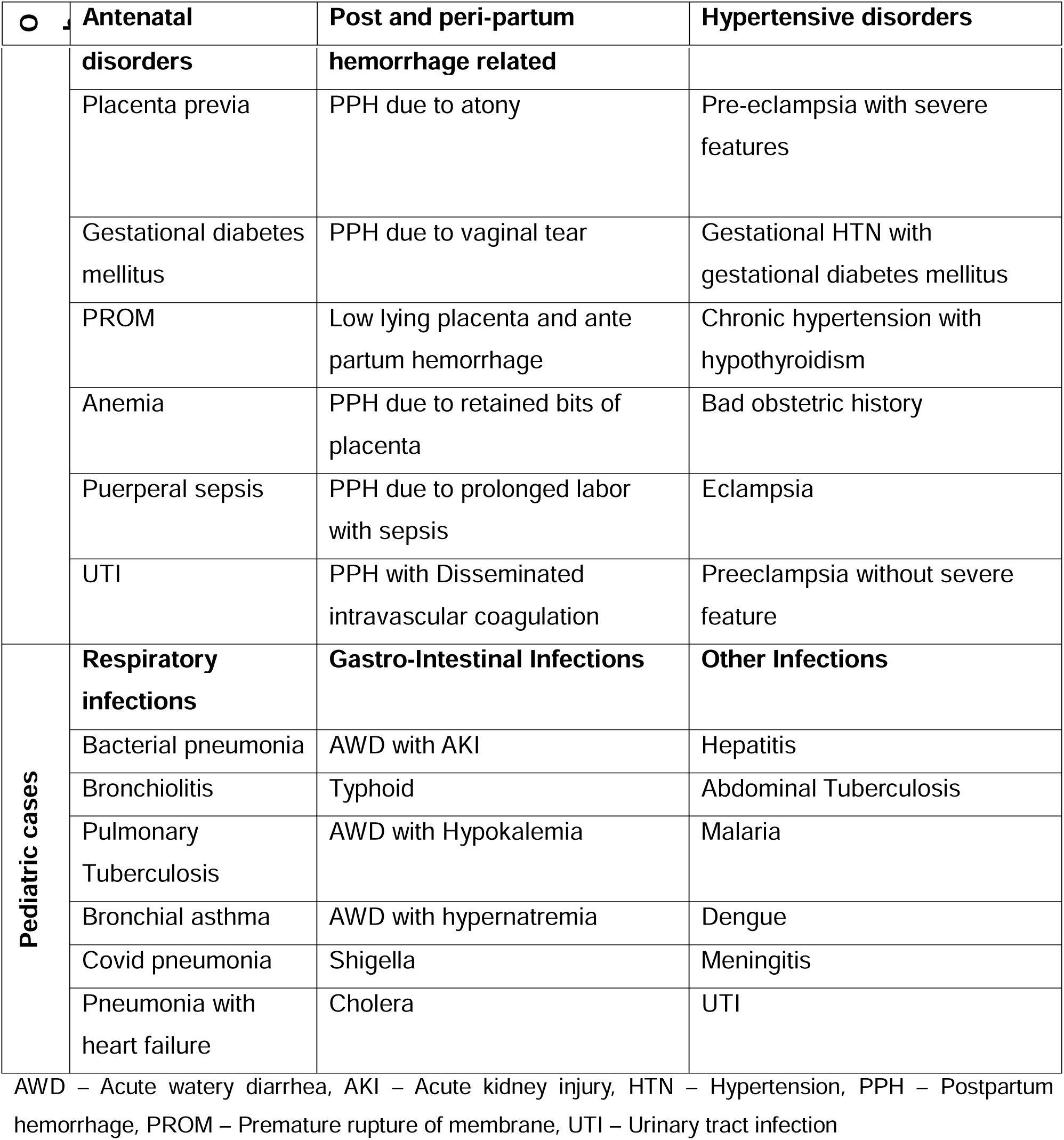
List of simulation cases of Obstetrics and Pediatrics.

| ○ | Antenatal | Post and peri-partum | Hypertensive disorders |
| --- | --- | --- | --- |
|  | **disorders** | **hemorrhage related** |  |
|  | Placenta previa | PPH due to atony | Pre-eclampsia with severe features |
|  | Gestational diabetes mellitus | PPH due to vaginal tear | Gestational HTN with gestational diabetes mellitus |
|  | PROM | Low lying placenta and ante partum hemorrhage | Chronic hypertension with hypothyroidism |
|  | Anemia | PPH due to retained bits of placenta | Bad obstetric history |
|  | Puerperal sepsis | PPH due to prolonged labor with sepsis | Eclampsia |
|  | UTI | PPH with Disseminated intravascular coagulation | Preeclampsia without severe feature |
| **Pediatric cases** | **Respiratory infections** | **Gastro-Intestinal Infections** | **Other Infections** |
|  | Bacterial pneumonia | AWD with AKI | Hepatitis |
|  | Bronchiolitis | Typhoid | Abdominal Tuberculosis |
|  | Pulmonary Tuberculosis | AWD with Hypokalemia | Malaria |
|  | Bronchial asthma | AWD with hypernatremia | Dengue |
|  | Covid pneumonia | Shigella | Meningitis |
|  | Pneumonia with heart failure | Cholera | UTI |

AWD – Acute watery diarrhea, AKI – Acute kidney injury, HTN – Hypertension, PPH – Postpartum hemorrhage, PROM – Premature rupture of membrane, UTI – Urinary tract infection

#### Formation of an expert team

Two dedicated case-writing teams have been established, one for Obstetrics and Gynecology care and another for Pediatric care. A senior expert clinician leads each team in their field. They developed simulated cases for the study. They conducted in-depth literature reviews to gather evidence on current clinical practices, as well as national and international guidelines, and available tools. The teams meet regularly to review the cases and ensure they follow current evidence-based guidelines and standard protocols that fit local practice.

#### Developing simulated cases using the EQIS platform

The EQIS platform is designed to facilitate efficient and structured case writing. It is structured as case for assessments, with embedded, real-time, evidence-based feedback. Each case is prepared as a physician-patient interaction scenario where a simulated patient presents to the physician with health-related complaints and is then assessed, diagnosed, and managed by the physician just as in a real clinical environment.

The case-development process had several phases. First, an expert case-writer drafted each case. Next, a lead expert clinician reviewed it. Once finalized, the cases were uploaded to the EQIS platform. Each case follows a structured case format across six domains – history taking, examination, investigations, diagnosis, treatment, and follow-up care – with 10-12 options in each domain. After the expert case-writer drafted the cases, AI-assisted technology (ChatGPT) created the initial skeleton for the cases to ensure consistency in formatting. After initial drafting and AI input, the expert clinicians reviewed and finalized the structured cases to ensure content alignment with standard guidelines and adaptation to local practice. Once completed each case followed a specific format and represented simulated clinical scenarios covering all six domains of patient care.

#### Enrollment of participating physicians

The research team works with hospital authorities to identify and develop an approach that enrolled all eligible participating physicians. An electronic register was created to record physician information and contact details – including email addresses and phone numbers –in a structured database. The dataset was regularly reviewed and updated before each round to include newly joined physicians and exclude those who leave the hospital, transfer to different departments, or retire from services. All physician information (including case scores) is confidential.

#### Physicians’ access to the EQIS platform

An individual account was created for each participant using their contact details. The research team sent the EQIS platform access link through email or other messaging services like WhatsApp. Participants logged in using their email ID and a one-time password (OTP).

#### Baseline survey

Prior to the initial round of data collection, baseline participant information will be collected at the individual level. This will include professional designation, educational qualifications, and years of experience. These details will be integrated into the EQIS platform and will appear upon the participant’s first login. The survey will be completed before accessing their assigned cases. In addition, facility-level information on staffing and available infrastructure will be obtained from the Heads of Departments at participating hospitals.

#### Training and orientation

All participants will be familiarized with the EQIS platform before data collection begins. The platform will be introduced through hospital-based orientation sessions using PowerPoint presentations, during which physicians will receive training on its use.

#### Case allocation

The cases will be allocated in groups of 3 to the participating physicians. Performance will be evaluated over six rounds during a 24-month period, with four-month gaps between rounds. Participants will care for simulated cases by making clinical decisions related to patient management. Their decisions will be scored against explicit, evidence-based criteria and compared with those of their peers through an inbuilt scoring system. They will be able to complete their assigned cases at their convenience within a defined period. Cases will be accessible via computer, tablet, or phone with an internet connection. Physicians will complete three cases in each round. Each case-completion will take about 20 minutes.

#### Feedback and progress monitoring

After completing each case, this platform will provide individual scores anonymously, promoting a safe platform for learning and self-improvement. This will appear on the individual participant’s EQIS dashboard. The platform will also compare group scores from round to round to monitor progress (Figure 3). As participants progress through each case, they will receive individualized feedback with practical clinical advice. The feedback will be categorized as necessary/correct, unnecessary/incorrect, or areas for standardization/improvement. Up-to-date, evidence-based references will support all feedback.

**Figure 3:**
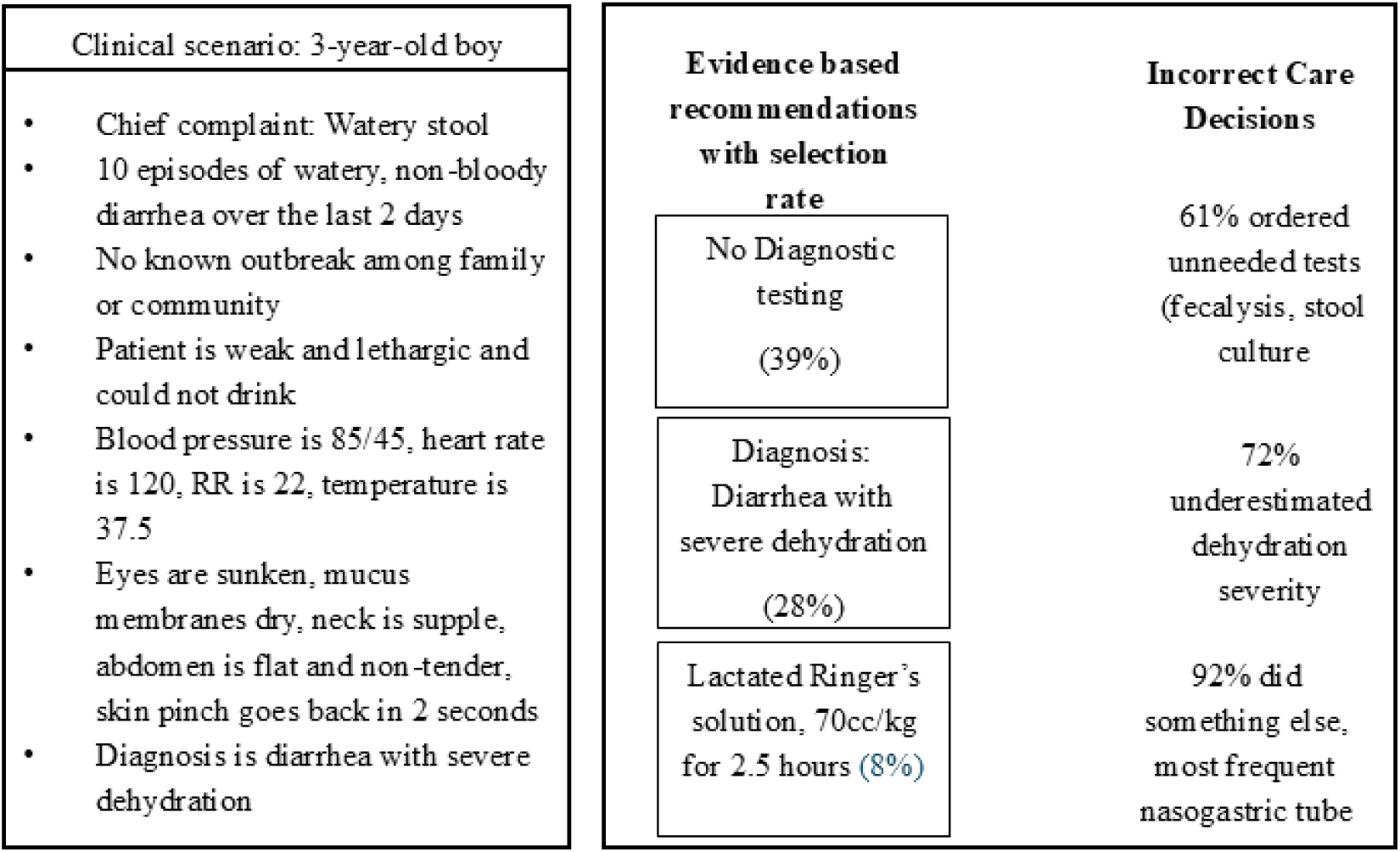
An example of group data output on the EQIS dashboard.

#### Evaluating performance

Each of the six cases for each of the three clinical topics includes evidence-based performance criteria listed on the participant scoring sheet. Responses from each participant will be scored against these explicit criteria to determine individual (confidential) performance. These individual scores are a key component of data collection, providing both real-time and post-round feedback, which is delivered anonymously to each participant. The data analytics team will aggregate participant responses to generate summary metrics (e.g., mean performance, score distributions), which will be used to provide insights to both individual participants and institutional leadership (Figure 4).

**Figure 4:**
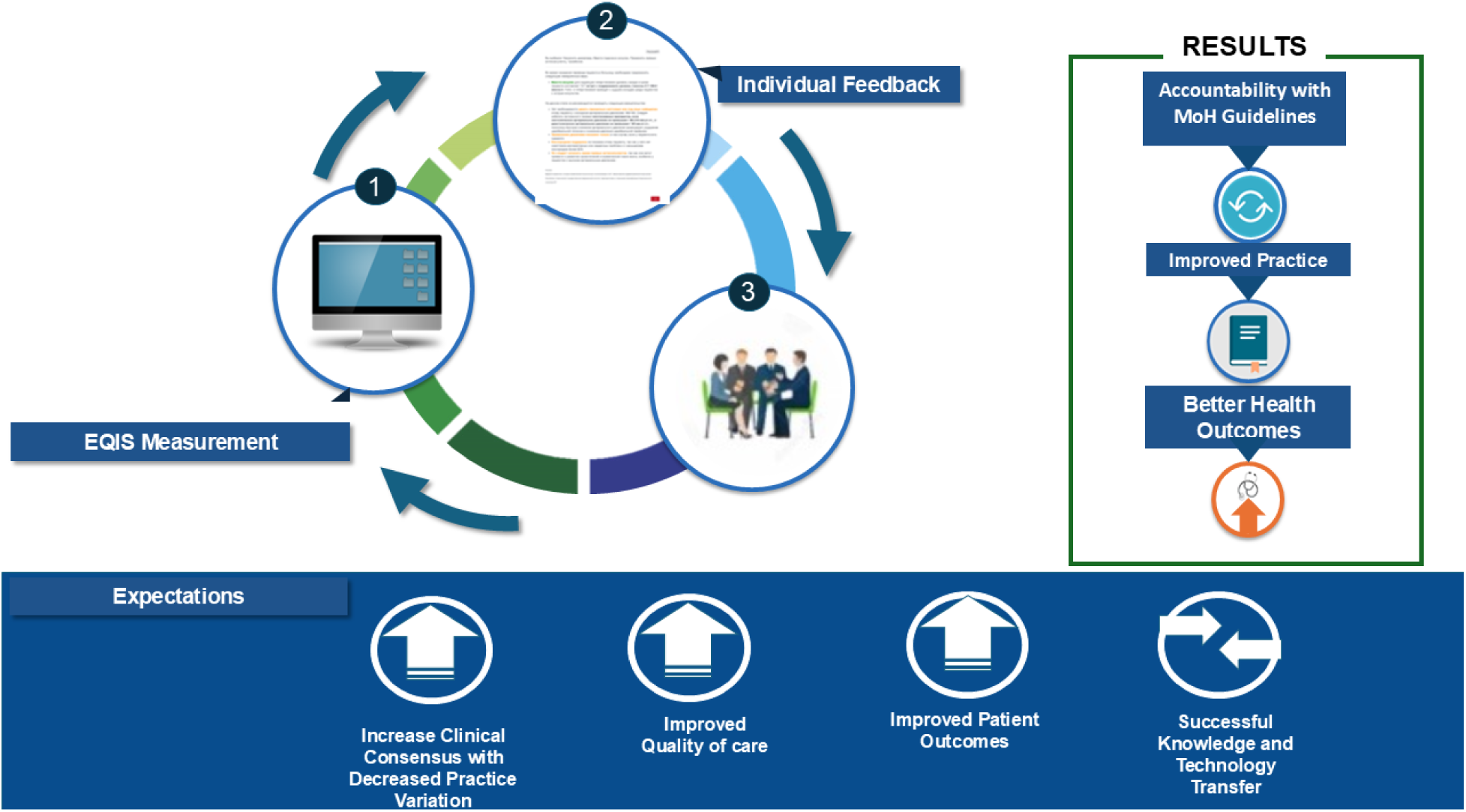
Individual and group feedback from the EQIS platform and expected results.

### Pre-post impact assessment

A pre-impact assessment will be conducted among patients attending the participating hospitals before the first round of the EQIS platform is initiated. This assessment will focus on key indicators, including patient satisfaction, cost of care, referral patterns, and facility utilization. Following the completion of the final round of the EQIS platform application, a post-impact assessment will be carried out in the same hospitals. The objective of pre- and post-impact assessment will be to examine the impact of implementing the EQIS platform on patient satisfaction, healthcare costs, referral patterns, and facility utilization. This will involve patients admitted to the departments of maternal and pediatric units. Eligible participants will include women aged 18 years or older admitted to the maternal department, as well as parents or legal guardians of children aged 1-59 months admitted to the pediatric department. Patients will be excluded if they are severely ill and unable to participate in the study activities or if they or their caregivers decline to provide informed consent.

### Data Analysis

We will evaluate the performance trends of the physicians. Descriptive statistics (i.e., means, standard deviations, and proportions) will summarize participation and completion rates along with overall quality scores across different care areas like clinical work-up, diagnosis, and treatment. To evaluate learning progress, repeated measures analysis, such as linear mixed-effects models or generalized estimating equations, will be used to assess score trends over the six rounds. We will track performance on key learning objectives identified at baseline. Subgroup analyses will be conducted to examine differences by gender, professional group, and hospital. We will use independent t-tests, ANOVA, or non-parametric equivalents, such as Mann-Whitney U or Kruskal-Wallis tests, when appropriate. To compare categorical variables, we will use chi-square or Fisher’s exact tests.

For the pre- and post-impact assessments, we will carry out comparative analyses to investigate differences in patient satisfaction, cost of care, referral patterns, and facility utilization. T-tests or Wilcoxon signed-rank tests for continuous outcomes will be used. Chi-square or Fisher’s exact tests will be applied for categorical outcomes. Multivariate regression models will be used to control potential confounders and to identify predictors of improved outcomes.

Statistical software such as Stata or R will be used for all statistical analyses. A p-value <0.05 will be considered statistically significant.

### Data Storage

Data will be entered via a web-based entry system and stored on a secure cloud server. Pre-and post-assessment data will be stored on a password-protected SQL Server 2008 R2 located in Dhaka, Bangladesh. All tablets and servers used in this project will be password protected.

### Ethics and Dissemination

Ethical approval was obtained from the National Research Ethics Committee of Bangladesh Medical Research Council, Bangladesh (Registration Number: 59524062024). For the simulated patient cases, informed written consent will be obtained from all participants. For the pre- and post-assessment, informed written consent will be obtained from all participants or guardians (for child participants). All methods were performed in accordance with the relevant guidelines and regulations, including the Declaration of Helsinki.

The study findings will be shared with participants at each hospital through meetings. In addition, a national-level dissemination meeting will be organized with the Ministry of Health and other stakeholders. The results will be put together in a report that includes an overview of the research objectives, methods, findings, and conclusions. Based on these findings, we will propose recommendations to improve the quality of clinical performance and standardize clinical practices among physicians to enhance maternal and pediatric care in Bangladesh. We will share the results through scientific articles, conference presentations, and distribution to relevant stakeholders.

## DISCUSSION

This study protocol outlines the use of the EQIS platform, a simulation-based platform. To our knowledge, this is the first study in Bangladesh that will use simulated clinical cases to evaluate and improve the performance of doctors in Obstetrics, Gynecology, and Pediatrics. By including scenarios that follow both national and international guidelines, the EQIS platform will provide a safe space for physicians to practice a variety of cases. This method aims to help them develop their critical thinking skills and receive structured feedback.

We intend to introduce the EQIS platform in Bangladesh to address inconsistencies in clinical practice that may compromise maternal and child health outcomes. The platform will encourage reflection, self-evaluation, and continuous learning by exposing physicians to standardized guideline-based cases. This will test the performance of the tool in the country, test the performances of the physicians and need of improvement and possible local data to allow stake-holders decision making process. This structured approach is especially important in LMICs, where physicians have fewer training opportunities, and building their skills is key to improving the health system.

If this study is successful, it could greatly impact clinical training and regulation in Bangladesh and similar settings in other LMICs. The EQIS platform can support healthcare organizations and professional groups in monitoring and improving competency standards, unifying clinical practices, and promoting accountability. Beyond its current focus, the platform could expand to include rural facilities, be tailored for other specialties and fit into ongoing medical education systems. We plan to introduce the EQIS platform to regulatory authorities, once substantiated by this study, to use for licensing and re-licensing in Bangladesh. The lessons gained from this study will be important for making the approach sustainable and supporting its wider use across healthcare settings.

## Data Availability

Not applicable (this is a protocol paper).

## Acknowledgements

We are grateful for the support and contributions of the participating hospitals and the Ministry of Health and Family Welfare, Government of Bangladesh. We are also grateful to Anjuman Ara Rita, Taslima Akter, Runa Akhter Dola, Ismat Jahan, Sharmin Afroze, Maimuna Sayeed, Sharika Nuzhat for their contribution in developing the clinical cases.

## DECLARATIONS

### Contributors

SA and JP conceptualized and designed this study. AHB, AMK and CAK critically reviewed the study design. FA, SBI, SJ, and MMH are involved in project management. SJ, NHC and MSI will be responsible for data management. SA, AMK and SBI drafted the manuscript, and all authors critically reviewed and approved the final manuscript before submission.

### Patient and public involvement

The research team engaged extensively with professional bodies, the Ministry of Health, hospital authorities and relevant stakeholders to ensure the feasibility and contextual relevance of the study. The study protocol was discussed with expert obstetricians and pediatricians, representatives of the Ministry of Health, hospital administrators, and treating physicians. Separate meetings were conducted at participating hospitals to review operational aspects. These consultations informed refinement of the study design and implementation strategy.

### Patient consent for publication

Not applicable.

